# Stalled Outcome Gains Despite Treatment Intensification in Stroke Patients Aged 80 and Older: A 12-Year Cohort Study

**DOI:** 10.64898/2026.07.23.26358827

**Authors:** Jonguk Kim, Do Yeon Kim, Nakhoon Kim, Jun Yup Kim, Jihoon Kang, Beom Joon Kim, Moon-Ku Han, Tai Hwan Park, Keon-Joo Lee, Joon-Tae Kim, Kang-Ho Choi, Jong-Moo Park, Kyusik Kang, Soo Joo Lee, Jae Guk Kim, Jae-Kwan Cha, Dae-Hyun Kim, Kyungbok Lee, Jeong-Yoon Lee, Jun Lee, Keun-Sik Hong, Yong-Jin Cho, Hong-Kyun Park, Byung-Chul Lee, Kyung-Ho Yu, Minwoo Lee, Dong-Eog Kim, Jay Chol Choi, Jee-Hyun Kwon, Wook-Joo Kim, Dong-Ick Shin, Kyu Sun Yum, Sung Il Sohn, Jeong-ho Hong, Sang-Hwa Lee, Ji Sung Lee, Juneyoung Lee, Philip B. Gorelick, Hee-Joon Bae, the CRCS-K Investigators

**Affiliations:** Department of Neurology, Seoul National University Bundang Hospital, Seongnam, Korea; Department of Neurology, Seoul Medical Center, Seoul, Korea; Department of Neurology, Korea University Guro Hospital, Seoul, Korea; Department of Neurology, Chonnam National University Hospital, Gwangju, Korea; Department of Neurology, Uijeongbu Eulji Medical Center, Eulji University, Uijeongbu, Korea; Department of Neurology, Nowon Eulji Medical Center, Eulji University School of Medicine, Seoul, Korea; Department of Neurology, Eulji University Hospital, Daejeon, Korea; Department of Neurology, Dong-A University Hospital, Busan, Korea; Department of Neurology, Soonchunhyang University Hospital, Seoul, Korea; Department of Neurology, Yeungnam University Medical Center, Daegu, Korea; Department of Neurology, Inje University Ilsan Paik Hospital, Goyang, Korea; Department of Neurology, Hallym University Sacred Heart Hospital, Anyang, Korea; Department of Neurology, Dongguk University Ilsan Hospital, Goyang, Korea; Department of Neurology, Jeju National University Hospital, Jeju, Korea; Department of Neurology, Ulsan University Hospital, Ulsan, Korea; Department of Neurology, Chungbuk National University & Hospital, Cheongju, Korea; Department of Neurology, Keimyung University Dongsan Hospital, Daegu, Korea; Department of Neurology, Hallym University Chuncheon Sacred Heart Hospital, Chuncheon, Korea; Department of Neurology, Clinical Research Center, Asan Medical Center, Seoul, Korea; Department of Neurology, Department of Biostatistics, Korea University, Seoul, Korea; Department of Neurology, Davee Department of Neurology, Northwestern University Feinberg School of Medicine, Chicago, Illinois, USA

## Abstract

**Background:** Adults aged 80 years and older are the fastest-growing segment of the stroke population. Whether contemporary advances in stroke care have produced sustained outcome gains in this group during the past decade, including the post-pandemic era, has not been examined in nationwide longitudinal data with concurrent age-specific comparison.

**Methods:** Using the Clinical Research Collaboration for Stroke in Korea-National Institutes of Health (CRCS-K-NIH) registry, we analyzed consecutive adults with acute ischemic stroke admitted within 7 days of symptom onset between 2011 and 2022. Patients aged 80 years and older formed the primary cohort; those aged 18-79 years served as comparators. Primary outcomes were 3-month utility-weighted modified Rankin Scale (UW-mRS) and 1-year all-cause mortality. Non-linear trends and age-group-by-year interaction were modeled using restricted cubic splines adjusted for sex, age, and baseline NIHSS.

**Results:** Of 83,953 patients, 18,514 (22.1%) were aged 80 years and older; their proportion rose from 17.4% to 27.3%. Despite treatment intensification (endovascular thrombectomy 4.2% to 10.6%; oral anticoagulation for atrial fibrillation 52.5% to 76.9%), 1-year mortality in this group fell from 27.5% in 2011 to 19.4% in 2019, then rose to 23.7% by 2022; 3-month UW-mRS improved from 0.47 to 0.51 over the same period, then stagnated at 0.48. Restricted cubic splines confirmed significant nonlinearity in both outcomes after adjustment for sex, age, and NIHSS (P < 0.01 for both). Younger patients showed no comparable functional deterioration. Age-group-by-year interaction was significant for 3-month UW-mRS (P < 0.001) but not for 1-year mortality (P = 0.058), which showed a similar direction in both age groups.

**Conclusion:** In this nationwide registry, patients aged 80 years and older experienced an age-specific inflection of functional recovery gains after 2019 despite continued intensification of acute stroke care. Sustaining outcome gains in this population may require attention to post-acute care infrastructure.

## Introduction

Adults aged 80 years and older, the “oldest-old”, are the fastest-growing demographic group globally ^1,2^ and account for more than one in four acute ischemic strokes in many high-income settings.^3^ Compared with younger stroke patients, the oldest-old have higher case fatality, more severe residual disability, and greater reliance on long-term institutional care, with correspondingly higher healthcare costs.^4–6^ Despite their disproportionate share of stroke burden, oldest-old patients remain underrepresented in the major randomized trials that guide contemporary stroke management.^7,8^

Over the past decade, acute stroke management has changed substantially with the widespread implementation of endovascular thrombectomy (EVT), the adoption of dual antiplatelet therapy (DAPT), and the transition from warfarin to direct oral anticoagulants (DOACs) for atrial fibrillation.^9,10^ Although enrollment in clinical trials was limited, subgroup and observational data indicated that these treatments also benefit older patients.^11,12^

However, whether these therapeutic advances have reached the oldest-old in routine practice has been assessed primarily for reperfusion therapies^6,13^, and data on broader measures of acute treatment and secondary prevention remain limited. Moreover, whether such treatment intensification has translated into improved functional outcomes or long-term prognosis in this group remains largely unknown. This question is further complicated at times of system-level disruption in care, such as the COVID-19 pandemic.^14^ Although a deterioration in stroke care during the pandemic has been reported,^15–17^ to our knowledge, it remains uncertain whether the treatment patterns and outcomes for the oldest-old differed from those of younger counterparts during this period.

Using a nationwide multicenter stroke registry covering 12 years (2011-2022), we aimed to (1) evaluate trends in acute treatment, secondary prevention, and clinical outcomes among the oldest-old; (2) determine whether outcome improvements were sustained over the full study period, with a prespecified focus on the 2019-2020 transition given the COVID-19 pandemic onset; and (3) compare these trends with those of younger counterparts to determine whether age-specific disparities were present.

## Methods

### Study Design and Population

This study used data from the Clinical Research Collaboration for Stroke in Korea-National Institutes of Health (CRCS-K-NIH) registry.^18,19^ Established in 2008, CRCS-K-NIH prospectively collects detailed data on patients hospitalized for acute stroke or transient ischemic attack; by 2022, 19 centers were participating. We identified consecutive adults (≥18 years) admitted with acute ischemic stroke between January 2011 and December 2022, defined as cerebral ischemic symptoms lasting >24 hours or the presence of a corresponding diffusion-weighted MRI lesion. We included patients presenting within 7 days of symptom onset (first recognition of acute neurological symptoms). The primary analytic cohort comprised patients aged ≥80 years. Patients aged 18-79 years served as a comparator cohort for age-stratified analyses. The patient selection flowchart is provided in eFigure 1.

### Ethics Statement

Data collection within the CRCS-K-NIH registry was approved by the local institutional review boards (IRBs) of all participating centers. The secondary use of fully anonymized, pre-existing registry data for this analysis was approved by the IRB of Seoul National University Bundang Hospital (approval number B-2505-973-101) and other participating centers, which waived the requirement for individual informed consent given the retrospective, non-interventional nature of the analysis and the absence of identifiable patient information.

### Data Collection and Outcomes

Demographic characteristics, vascular risk factors, pre-stroke disability (modified Rankin Scale [mRS]), initial stroke severity (National Institutes of Health Stroke Scale [NIHSS]), stroke subtype according to the Trial of Org 10172 in Acute Stroke Treatment (TOAST) criteria with modifications,^20^ time metrics including symptom onset time and last-known-well time, and administration of intravenous thrombolysis (IVT), EVT, and discharge medications were extracted from the registry database. For vascular risk factors, treatment status before stroke onset was classified as “on medication” or “not on medication”. Inpatient interdepartmental transfers were recorded, with transfers to rehabilitation facilities being systematically documented from 2017 onward, and discharge destination was categorized as home or institutional care.

Information on clinical outcomes was prospectively obtained through structured telephone interviews or outpatient assessments at 3 months and 1 year after stroke onset, following standardized protocols. The primary outcomes were 3-month functional status assessed by utility-weighted mRS (UW-mRS) and 1-year all-cause mortality. UW-mRS assigns preference-based utility weights to each mRS grade (ranging from 0 for death to 1 for no disability), converting the ordinal scale into a continuous measure that captures the full spectrum of post-stroke disability.^21^ Secondary outcomes included the proportion achieving mRS 0-2 at 3 months, recurrent stroke at 1 year, and a composite vascular outcome comprising recurrent stroke, acute myocardial infarction, or death at 1 year.

### Statistical Analysis

Descriptive analyses summarized clinical characteristics, treatment, and outcomes by calendar year (2011-2022). Temporal trends were evaluated using linear contrasts for continuous variables and the Cochran-Armitage test for categorical variables. Outcomes were analyzed using linear (UW-mRS), logistic (mRS 0-2) and Cox regression (recurrent stroke, all-cause mortality, and composite vascular events, with 365-day cumulative incidence estimated using Kaplan-Meier methods). Within-hospital clustering was accounted for using generalized estimating equations for the linear and logistic models and cluster-robust (sandwich) standard errors for the Cox models.

For each outcome, we obtained year-specific estimates and regressed these annual estimates on calendar year as a continuous variable. Trends were assessed with two prespecified models: a crude model, providing the primary population-level description, and a parsimonious model adjusted for sex, age, and baseline NIHSS. The restricted cubic spline models were fitted at the individual patient level; the year-specific estimates and their regression on calendar year were used to describe and display the population-level annual trajectory. We adjusted only for these non-modifiable case-mix factors, because other characteristics may lie on the causal pathway of care improvement and would attenuate the trends of interest.

To assess whether there were changes in outcome trends around the COVID-19 pandemic onset, we fitted restricted cubic spline (RCS) functions with prespecified knots at 2013, 2019, and 2022. The sex, age, and NIHSS adjusted (parsimonious) RCS model was the main analysis. Age-specificity was examined by testing age-group-by-year interaction terms, derived from the RCS models, in the overall dataset. These analyses were restricted to the primary outcomes (3-month UW-mRS and 1-year all-cause mortality) to limit multiplicity.

Missing values were handled using pairwise deletion for bivariate analyses. In multivariable models, missing covariates were imputed using year-specific medians (continuous) or modes (categorical). For mRS, missing 3-month values were imputed as 6 if death occurred within 5 months; otherwise, 1-year mRS or discharge mRS was used. Missing 1-year mRS was imputed as 6 if death occurred within 15 months.

We conducted three prespecified sensitivity analyses. First, the RCS model was fitted with full adjustment, additionally including vascular risk factors (hypertension, diabetes, hyperlipidemia, atrial fibrillation, and smoking), pre-stroke mRS, and TOAST subtype, as a robustness check. Second, the primary analysis was repeated in the 12 centers that enrolled patients continuously throughout 2011-2022, given the increasing number of participating centers over time. Third, for the functional outcome, the analysis was restricted to complete cases with an observed 3-month UW-mRS.

We performed two post hoc exploratory analyses to assess potential contributors to the outcome changes during the COVID-19 period. First, because the overall rise in EVT use could mask reduced access among treatable patients, we examined EVT use by age group in thrombectomy-eligible patients (NIHSS ≥6)^22^ and severe strokes (NIHSS ≥14). Second, because 3-month functional outcome and 1-year mortality cover different follow-up windows, and because early and late post-stroke mortality have distinct determinants,^23^ we decomposed 1-year all-cause mortality into early (0-90 days) and late (90-365 days, conditional on 90-day survival) death and tested the age-group-by-year interaction for each.

All tests were two-sided (P < 0.05). Analyses used SAS 9.4 (SAS Institute Inc., Cary, NC, USA) and R 4.2.1 (R Foundation for Statistical Computing, Vienna, Austria), and reporting followed the Strengthening the Reporting of Observational Studies in Epidemiology (STROBE) statement.^24^

## Results

### Baseline Characteristics and Demographic Shifts

Between 2011 and 2022, a total of 83,953 acute ischemic stroke patients were enrolled in the CRCS-K-NIH registry. The average age of the total cohort was 67.8 ± 12.7 years, and 42.4% (n = 35,595) were women. Among these, 18,514 patients were classified as oldest-old (aged ≥80 years). The proportion of oldest-old stroke patients rose steadily from 17.4% in 2011 to 27.3% in 2022 (P for trend < 0.001).

The clinical phenotype within the oldest-old group also changed substantially over the study decade (Table 1). The prevalence of prior stroke and hypertension remained stable, whereas diabetes increased gradually and hyperlipidemia exhibited a U-shaped pattern with a nadir around 2014. Current smoking rates decreased by nearly half, and mean BMI rose slightly. Atrial fibrillation prevalence remained largely stable, while pre-stroke anticoagulation among patients with atrial fibrillation increased nearly threefold (11.7% to 33.5%). Over the same period, the proportion of cardioembolic stroke decreased from 32.3% to 27.7%. The proportion of oldest-old patients admitted via the emergency department increased significantly, but onset-to-hospital time remained largely unchanged throughout the study period.

**Table 1.**
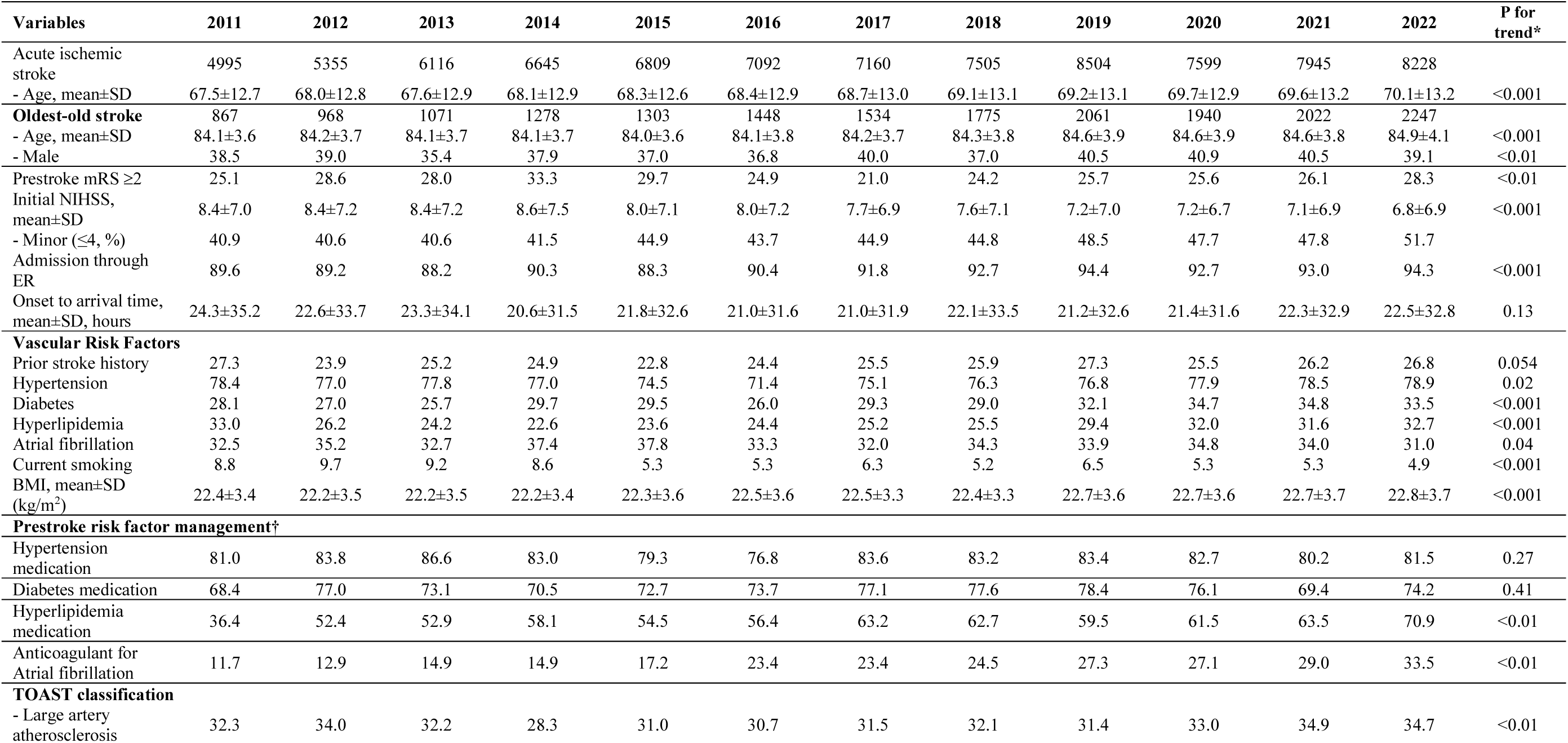
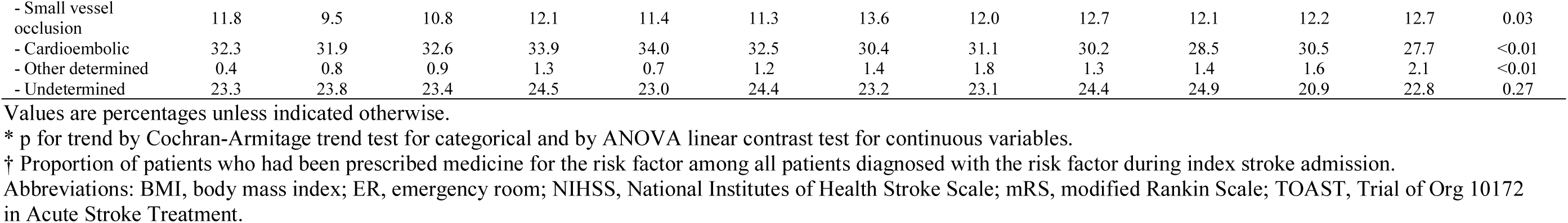
Temporal Trends in Baseline Characteristics of Patients Aged ≥80 Years with Acute Ischemic Stroke, 2011-2022.

### Treatment Trends in the Oldest-Old

Most aspects of stroke management in the oldest-old group improved significantly during the study decade (Table 2; Figure 1). The use of EVT increased approximately 2.5-fold, from 4.2% in 2011 to 10.6% in 2022. The use of DAPT during the acute phase increased from 24.1% to 44.2%. Among patients with atrial fibrillation, the prescription rate of oral anticoagulants at discharge, driven largely by the introduction of DOACs, rose significantly from 52.5% in 2011 to 76.9% in 2022. Statin prescription rates rose from 69.3% to 87.7% overall and exceeded 94% in patients with atherosclerotic or small-vessel occlusive stroke. In contrast, IVT use rose from 2011 to 2014, declined sharply from 2015 to a nadir of 7.3% in 2016, partially rebounded thereafter, and decreased again by 2022. Workflow time metrics remained stable (door-to-needle time P for trend = 0.64; door-to-puncture time P for trend = 0.73).

**Table 2.** Temporal Trends in Acute Treatment and Secondary Prevention Among Patients Aged ≥80 Years with Acute Ischemic Stroke, 2011-2022.

| Variables | 2011 | 2012 | 2013 | 2014 | 2015 | 2016 | 2017 | 2018 | 2019 | 2020 | 2021 | 2022 | P for trend* |
| --- | --- | --- | --- | --- | --- | --- | --- | --- | --- | --- | --- | --- | --- |
| **Reperfusion therapy** |  |  |  |  |  |  |  |  |  |  |  |  |  |
| IV thrombolysis rate | 11.9 | 13.4 | 12.9 | 14.9 | 8.7 | 7.3 | 8.9 | 8.7 | 11.1 | 10.7 | 10.0 | 8.6 | <.001 |
| Endovascular thrombectomy rate | 4.2 | 4.2 | 6.5 | 7.0 | 7.6 | 9.0 | 10.2 | 11.0 | 11.3 | 11.6 | 11.3 | 10.6 | <.001 |
| Door-to-needle time, median (IQR) | 46 (36-55) | 43 (32-52) | 36 (29-46) | 37 (28-47) | 39 (28-53) | 39 (29-53) | 38 (28-53) | 40 (28-55) | 39 (28-51) | 39 (27-54) | 37 (27-50) | 41 (30-57) | 0.64 |
| - within 45min | 44.3 | 54.8 | 72.8 | 71.2 | 59 | 61.2 | 58.1 | 57.4 | 60.3 | 60.6 | 63.2 | 56.8 | 0.83 |
| Door-to-puncture time, median (IQR) | 123 (89-140) | 100 (80-135) | 105 (80-139) | 106 (85-126) | 98 (79-128) | 96 (74-123) | 94 (71-124) | 99 (78-127) | 101 (77-131) | 100 (74-147) | 99 (79-139) | 110 (86-143) | 0.73 |
| - within 90m | 25.7 | 37.8 | 30.2 | 31.4 | 41.5 | 44.2 | 45.3 | 40.1 | 39.1 | 41.9 | 35.7 | 27.5 | 0.38 |
| **Secondary Prevention** |  |  |  |  |  |  |  |  |  |  |  |  |  |
| Dual antiplatelet, overall | 24.1 | 24.8 | 24.1 | 26.8 | 27.9 | 27.2 | 31.6 | 35.6 | 38.7 | 39.6 | 44.3 | 44.2 | <.001 |
| Dual antiplatelet in minor LAA, SVO | 31.9 | 37.1 | 35.2 | 45.7 | 48.8 | 45.8 | 50.0 | 63.4 | 63.5 | 69.0 | 75.7 | 76.2 | <.001 |
| Anticoagulant, overall | 20.8 | 22.2 | 24.0 | 25.3 | 28.3 | 26.0 | 27.3 | 29.2 | 31.4 | 30.1 | 30.2 | 30.4 | <.001 |
| Anticoagulant in AF | 52.5 | 47.8 | 60.0 | 57.9 | 64.9 | 66.2 | 72.1 | 72.6 | 79.0 | 74.4 | 74.2 | 76.9 | <.001 |
| DOAC in AF | 0.0 | 0.0 | 4.6 | 3.8 | 31.4 | 55.2 | 64.4 | 66.5 | 76.1 | 71.6 | 72.3 | 74.1 | <.001 |
| Statin, overall | 69.3 | 75.7 | 80.2 | 80.7 | 81.1 | 80.0 | 84.6 | 85.6 | 88.8 | 87.0 | 86.3 | 87.7 | <.001 |
| Statin in LAA, SVO | 77.0 | 83.4 | 88.1 | 91.9 | 87.7 | 86.5 | 92.3 | 93.9 | 93.7 | 93.4 | 94.2 | 94.5 | <.001 |
| **Rehabilitation** |  |  |  |  |  |  |  |  |  |  |  |  |  |
| Inpatient interdepartmental transfer† | 16.8 | 17 | 16.4 | 17.9 | 21.4 | 22.7 | 20.4 | 18.7 | 15 | 18.1 | 21.4 | 21.1 | 0.01 |
| - Rehabilitation transfer† | ... | ... | ... | ... | ... | ... | 11.7 | 12.4 | 10.9 | 13 | 15.6 | 16.3 | <.001 |
| Discharge to institutional care‡ | 42.2 | 34.6 | 28.3 | 38.9 | 33.5 | 39.3 | 41.2 | 39.3 | 38 | 37.8 | 37.4 | 34.2 | 0.82 |

Values are percentages unless indicated otherwise.
\* p for trend by Cochran-Armitage trend test for categorical and by linear contrast test for continuous variables.
† Denominator: patients with discharge mRS < 6 (alive at discharge). Inpatient rehabilitation transfer was not systematically recorded before 2017; annual percentages for 2011-2016 are not reported.
‡ Denominator restricted to patients with discharge mRS < 6 who were formally discharged (excluding early discharge against medical advice and inpatient interdepartment transfer); the registry does not distinguish between facility subtypes.
Abbreviations: AF, atrial fibrillation; DOAC, direct oral anticoagulant; LAA, large artery atherosclerosis; SVO, small vessel occlusion.

**Figure 1.**
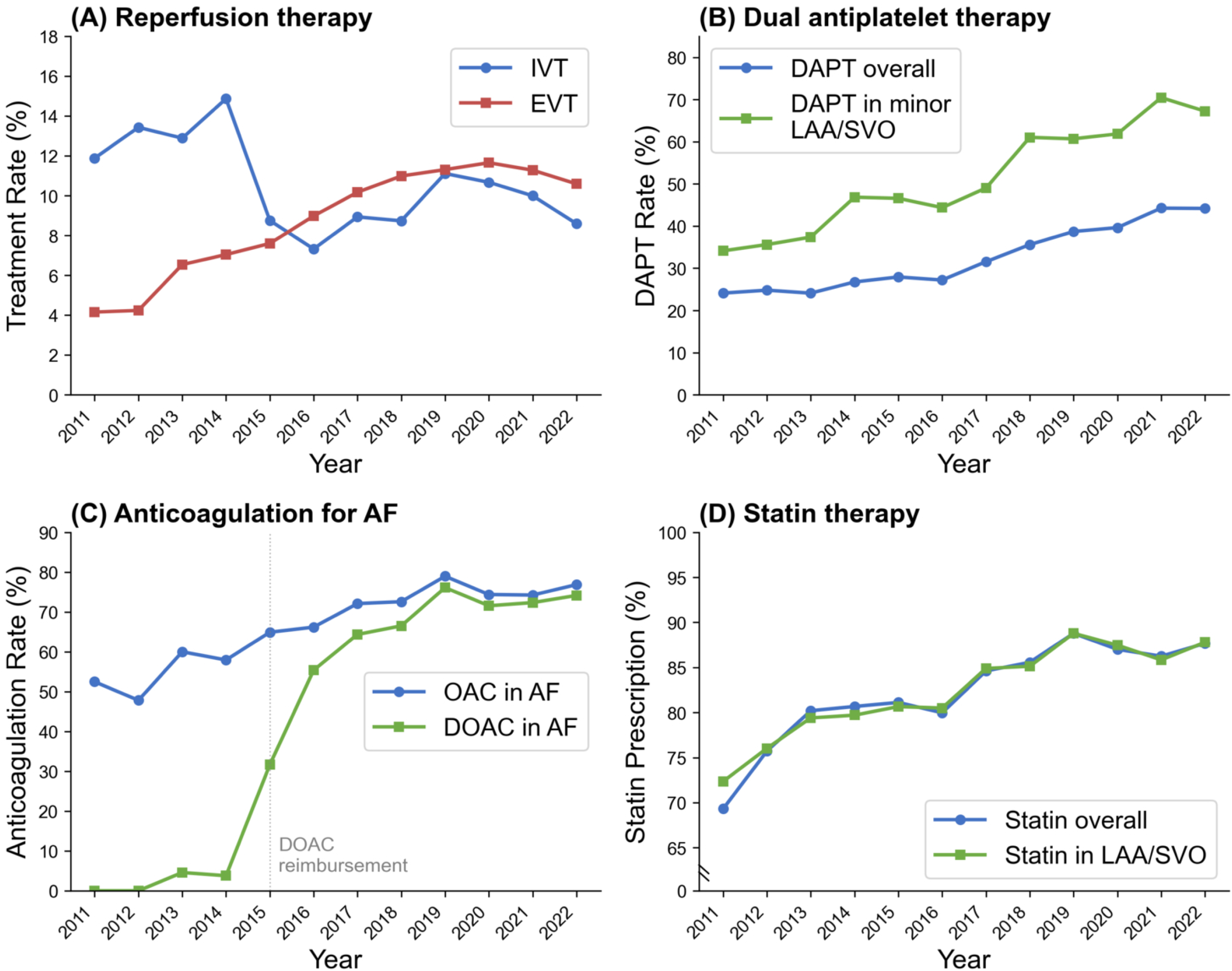
Temporal Trends in Stroke Management among Oldest-Old Patients, 2011-2022. Four panels display treatment patterns over time: (A) intravenous thrombolysis (IVT) and endovascular thrombectomy (EVT) rates; (B) dual antiplatelet therapy (DAPT) overall and in minor non-cardioembolic stroke (LAA/SVO subgroup); (C) oral anticoagulation (OAC) and direct oral anticoagulant (DOAC) use among patients with atrial fibrillation; (D) statin prescription overall and in atherosclerotic subtypes (LAA/SVO).

For post-acute care disposition, inpatient interdepartmental transfer and transfer to rehabilitation showed an overall increasing trend despite year-to-year fluctuation, while discharge to rehabilitation/long-term care facility remained stable.

### Outcome Trends in the Oldest-Old

Over the first nine years (2011-2019), clinical outcomes improved steadily (Table 3). The 1-year all-cause mortality rate declined from 27.5% in 2011 to 19.4% in 2019, and functional recovery, as measured by the 3-month UW-mRS, also improved during this period (from a mean of 0.47 ± 0.39 in 2011 to 0.51 ± 0.37 in 2019). After 2019, these gains were not sustained: 1-year mortality rose to 23.7%, whereas the mean 3-month UW-mRS plateaued at 0.48 from 2020 through 2022. The 4.3-percentage-point rise in 1-year mortality after 2019 offset more than half of the 8.1-percentage-point decline observed between 2011 and 2019. The proportion of patients achieving functional independence (3-month mRS 0-2) followed the same pattern, peaking near 2019 and declining thereafter. Other secondary outcomes followed similar patterns (eFigure 3).

**Table 3.** Temporal Trends in Clinical Outcomes Among Patients Aged ≥80 Years with Acute Ischemic Stroke, 2011-2022.

| Outcome Metric | 2011 | 2012 | 2013 | 2014 | 2015 | 2016 | 2017 | 2018 | 2019 | 2020 | 2021 | 2022 | Crude | P for trend*<br>Sex/Age/<br>NIHSS<br>Adjusted |
| --- | --- | --- | --- | --- | --- | --- | --- | --- | --- | --- | --- | --- | --- | --- |
| **3-month UW-mRS** | 0.47±0.39 | 0.45±0.38 | 0.47±0.38 | 0.47±0.38 | 0.49±0.37 | 0.48±0.38 | 0.49±0.37 | 0.50±0.37 | 0.51±0.37 | 0.48±0.37 | 0.48±0.37 | 0.48±0.38 | <0.01 | 0.17 |
| 3-month mRS 0-2 | 34.6 | 32.9 | 34.7 | 35.6 | 36.8 | 36.7 | 37.0 | 36.7 | 38.0 | 36.6 | 35.1 | 36.2 | 0.13 | 0.17 |
| **1-year stroke recurrence** | 6.7% | 6.6% | 4.8% | 4.5% | 6.6% | 4.3% | 5.3% | 4.6% | 5.2% | 5.9% | 5.8% | 6.0% | 0.67 | 0.68 |
| **1-year all-cause death** | 27.5% | 29.4% | 27.4% | 25.7% | 21.6% | 18.0% | 20.5% | 20.0% | 19.4% | 22.5% | 23.2% | 23.7% | 0.047 | 0.14 |
| **1-year composite event (recurrence, AMI, or death)** | 30.8% | 33.1% | 30.4% | 28.0% | 25.3% | 20.3% | 24.2% | 23.7% | 23.3% | 25.9% | 27.2% | 27.9% | 0.11 | 0.24 |

Values are mean ± SD for utility-weighted mRS (UW-mRS) and Kaplan-Meier estimates (%) for clinical events.
\* Trends in UW-mRS and mRS 0-2 were assessed using linear regression and logistic regression models, respectively. Time-to-event outcomes were analysed using marginal Cox proportional hazards models. All models accounted for hospital-level clustering.
Adjusted event probability by marginal Cox's PH regression with robust sandwich estimator & clustering for hospitals and P for linear trend by linear regression
Abbreviations: mRS, modified Rankin Scale; UW-mRS, utility-weighted mRS; NIHSS, National Institutes of Health Stroke Scale; AMI, acute myocardial infarction

The linear trends for 3-month UW-mRS and 1-year all-cause mortality were significant in crude analyses, but were attenuated after sex, age, and NIHSS adjustment (Table 3). Because the trajectory was visibly non-linear, a single linear slope cannot capture the full 12-year time course. To characterize these non-linear trends, we modeled non-linear trends with RCS. RCS models adjusted for sex, age, and NIHSS confirmed significant nonlinearity for both 3-month UW-mRS (P for nonlinearity = 0.007) and 1-year mortality (P for nonlinearity < 0.001; Figure 2).

**Figure 2.**
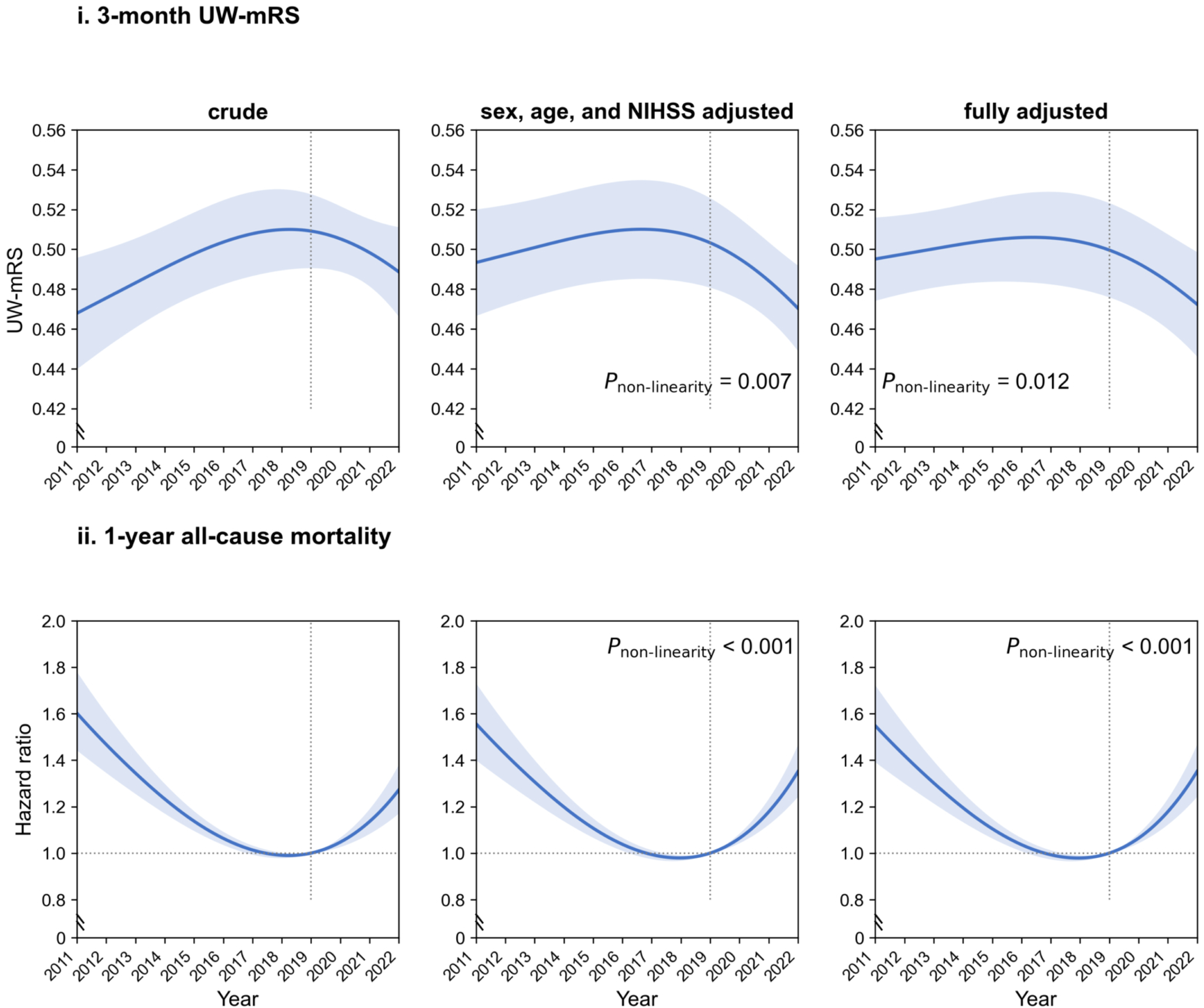
Restricted Cubic Spline Analysis of Temporal Outcome Trends among Oldest-Old Patients, 2011-2022. Restricted cubic spline curves (95% CI shaded; knots 2013, 2019, 2022) for oldest-old patients. Upper row, three-month UW-mRS (higher is better); lower row, 1-year all-cause mortality as a hazard ratio versus 2019. Columns: crude, parsimonious (sex, age, and NIHSS adjusted), and fully adjusted models. Nonlinearity P values are shown for the adjusted models.

### Comparison with Younger Patients

To determine whether the post-2019 inflection was specific to the oldest-old, we compared them with younger patients (18-79 years). The two groups differed across most baseline characteristics (eTables 1-2), and treatment intensified in both over the study period (eTable 3; eFigure 2).

In younger patients, both 3-month UW-mRS and 1-year all-cause mortality improved significantly in crude analyses, although these linear trends were no longer significant after sex, age, and NIHSS adjustment (eTable 4). For 1-year all-cause mortality, younger patients resembled the oldest-old, with an age-group-by-year interaction that was not statistically significant (P for interaction = 0.058; eTable 5). However, functional outcome was age-specific: unlike the oldest-old, younger patients showed no significant post-2019 inflection in 3-month UW-mRS (P for nonlinearity = 0.25), and the interaction confirmed this divergence (P for interaction < 0.001; Figure 3).

**Figure 3.**
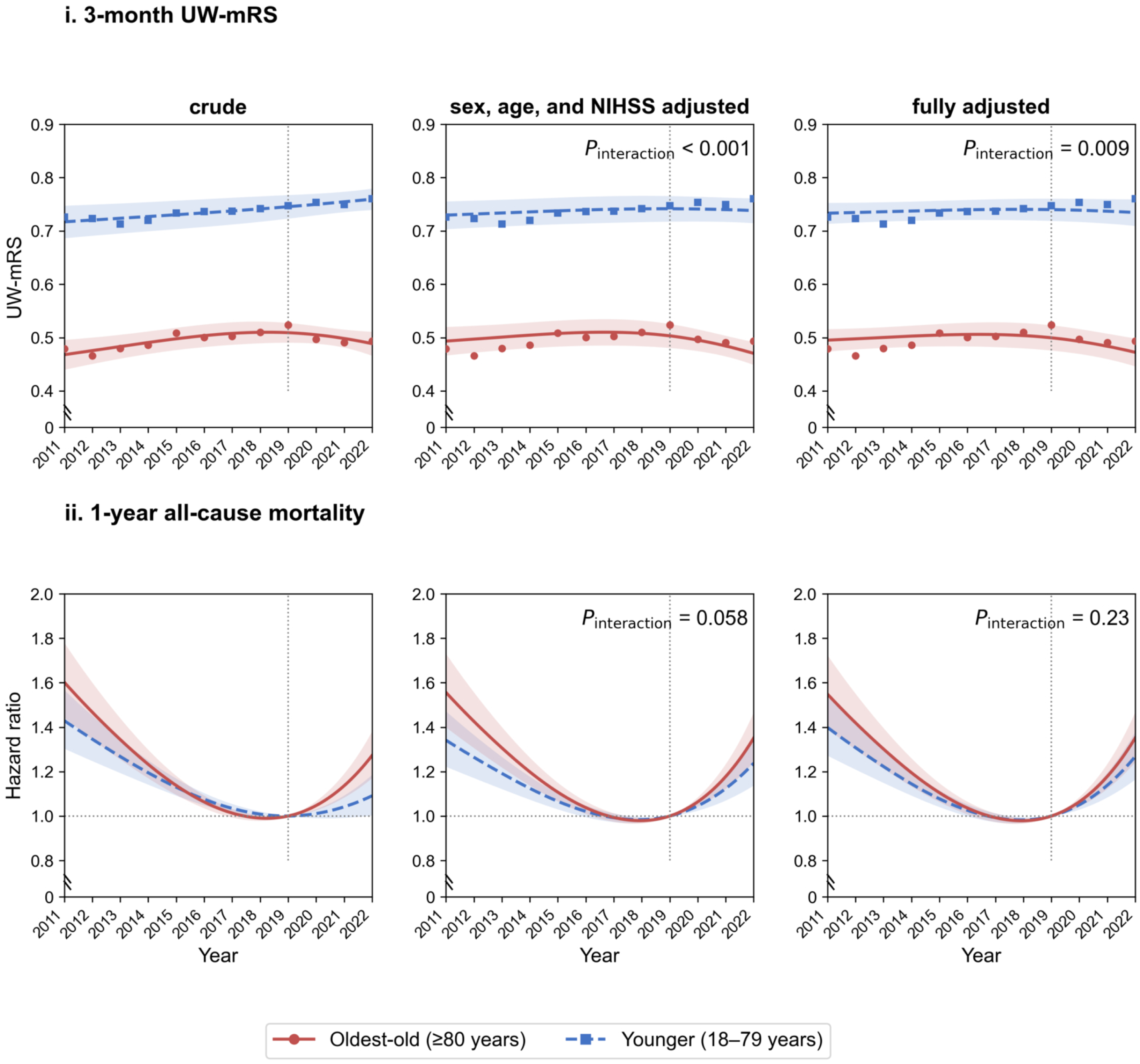
Age-Stratified Comparison of Functional Outcomes and Mortality, 2011-2022. Oldest-old (≥80 years; red solid) versus younger patients (18-79 years; blue dashed); points are observed annual mean UW-mRS. Restricted cubic spline curves (95% CI shaded; knots 2013, 2019, 2022). Upper row, three-month UW-mRS; low row, 1-year all-cause mortality as a hazard ratio versus 2019. Columns: crude, parsimonious (sex, age, and NIHSS adjusted), and fully adjusted models. Age-group-by-year interaction P values are shown for the adjusted models.

### Sensitivity Analyses

The age-specific divergence in 3-month UW-mRS was consistent across sensitivity analyses. In the fully adjusted model, the age-group-by-year interaction for 3-month UW-mRS remained significant (P for interaction < 0.001), whereas that for 1-year all-cause mortality was not (P for interaction = 0.17; eTable 5). This pattern, significant for functional outcome but not for mortality, also held in the 12 centers enrolling throughout 2011-2022 (eTable 6), and a complete-case analysis restricted to non-imputed 3-month mRS was concordant (eTable 7).

### Exploratory Analyses

Among potentially thrombectomy-eligible patients (NIHSS ≥6), EVT use in the oldest-old plateaued after 2019 (24.1% in 2019 to 22.9% in 2022), whereas it continued to rise in younger patients (30.1% to 31.2%), with a wider age gap in severe strokes (NIHSS ≥14; eTable 8). In the mortality decomposition, both early (10.6% to 12.7%) and late (10.4% to 11.9%) death rose after 2019 in the oldest-old, with little change in younger patients (early, 4.0% to 3.8%; late, 2.6% to 3.2%); the interaction was significant for early mortality (P for interaction < 0.001) but not for late mortality (P for interaction = 0.23; eTable 9).

## Discussion

To our knowledge, this is the first age-stratified study to track both 3-month function and 1-year mortality across the contemporary treatment and pandemic eras. In this 12-year nationwide registry study of 18,514 oldest-old stroke patients, evidence-based treatment intensified throughout the decade, with DAPT use doubling, anticoagulation for atrial fibrillation expanding substantially, and EVT increasing approximately 2.5-fold, yet outcomes did not track these gains. Nine years of steady improvement, during which 1-year mortality fell from 27.5% to 19.4% with concurrent functional gains, were followed by a post-2019 inflection, 1-year mortality rose to 23.7% and UW-mRS stagnated. This inflection was age-specific: the age-group-by-year interaction was significant for 3-month functional outcome but not statistically significant for 1-year mortality (P = 0.058).

The outcome trends we observed are concordant with previous reports. Before the pandemic, the Get With The Guidelines-Stroke program documented sustained in-hospital quality and outcome gains over two decades,^25^ and the Swedish Stroke Register reported improving functional outcome and declining case-fatality among older patients through 2019.^26^ The direction of treatment intensification we observed, including the rise in thrombectomy and the shift from warfarin to direct oral anticoagulants, matches these registries, indicating that the pre-2019 improvement was not specific to Korea. The post-2019 deterioration agrees with pandemic-era reports of worse acute stroke outcomes and higher in-hospital mortality among older patients.^27^ Within our cohort, the post-2019 change was consistent across 3-month function, 1-year mortality, and secondary outcomes, and persisted under full adjustment, restriction to the 12 continuously enrolling centers, and complete-case analysis.

The pre-2019 gains coincided with the wide adoption of evidence-based care, including EVT use that exceeded oldest-old rates in US registries^13^ and dual antiplatelet therapy after the CHANCE and POINT trials.^28–30^ This intensification was sustained after 2019, and in-hospital workflow was preserved, with stable onset-to-arrival and no worsening of door-to-needle or door-to-puncture times. However, although the overall EVT rate was sustained, among thrombectomy-eligible patients it edged down in the oldest-old while rising in younger patients, with the widest gap in severe strokes. IVT also declined around 2015, most likely from greater use of direct EVT and DOAC-related contraindications in this atrial fibrillation–rich population.^31–34^ Because acute treatment and workflow broadly were maintained, and because over 80% of the oldest-old received no reperfusion therapy, acute care is an unlikely primary cause of the post-2019 inflection.

One possibility is that the deterioration arose after discharge, where recovery in the oldest-old depends on rehabilitation and continuity of secondary prevention.^35,36^ Post-acute care volume, measured by rehabilitation transfers and institutional discharge, was maintained during 2020-2022. However, the registry does not capture the quality or intensity of that care, which several reports suggest declined during the pandemic.^37–39^ The inflection may thus reflect declining post-acute care quality rather than reduced access. Additional pandemic-era pressures may have compounded this, including weakened social and caregiving support,^40,41^ the attenuation of family-centered eldercare in Korea,^42,43^ direct effects of COVID-19 infection,^44^ and workforce strain in neurology and emergency medicine, documented internationally^45,46^ and pronounced in Korea.^47^ These mechanisms are not mutually exclusive, and our data cannot distinguish among them.

Although the age-specific signal was clearer for function than for mortality, this may reflect the sensitivity and timing of the endpoints rather than a true divergence. The UW-mRS is a continuous, granular measure that detects a modest effect more readily than all-cause mortality. Mortality is coarser and further diluted in the oldest-old by deaths unrelated to the index stroke.^36,48^ The timing of outcome tracking points the same way. When 1-year mortality was decomposed, the age-specific excess was concentrated in early death (0-90 days; P for interaction < 0.001), whereas the later increase (90-365 days) was similar in both age groups (P = 0.23). Because early post-stroke death is driven mainly by stroke severity and age, and later death additionally by cardiovascular comorbidity,^23^ the age-specific signal fell in the early, stroke-related window. The later increase observed in both groups was modest, and its cause cannot be determined from our data, though residual pandemic-related effects are possible. The age-specific vulnerability was therefore most evident in functional outcome and early mortality. However, the mortality comparisons warrant some caution, as deaths were far less frequent in younger patients.

Our findings have implications for how stroke care quality is monitored in older patients. Because the oldest-old deteriorated while acute treatment and workflow were maintained, acute process metrics alone can miss where quality is slipping.^25^ The post-acute and community-care capacity most likely underlying the inflection is not routinely captured by stroke registries. Monitoring in this group should therefore track the specific domains implicated here, including reperfusion access among eligible and severe strokes, the quality and intensity rather than the volume of post-acute rehabilitation, and community and caregiving support.^49,40,41^ Prospective evaluation of these domains is needed to confirm their contribution and to guide targeted quality improvement.

Several features of stroke presentation in the oldest-old also changed over the period. The decline in cardioembolic stroke despite stable atrial fibrillation prevalence, alongside increased pre-stroke anticoagulation, suggests improved primary prevention. Declining NIHSS scores and more frequent minor stroke presentations likely reflect improved detection.^1^ Diabetes prevalence rose in the oldest-old but not in younger patients,^50^ possibly reflecting cohort effects of economic transition and dietary Westernization, as well as statin-associated incident diabetes.^51^

This study has several limitations. First, unmeasured confounders, including pandemic-related changes in frailty and healthcare-seeking behavior, cannot be excluded. Second, although proxy measures of post-acute care use were examined, we did not capture rehabilitation intensity, outpatient adherence, or caregiving quality, the domains most likely involved. Third, the 2019 knot used to model the trend change was prespecified from the known pandemic onset rather than identified by formal changepoint detection. Fourth, the landmark and thrombectomy analyses were exploratory and hypothesis-generating. Fifth, the number of participating centers increased over the study period. The main analyses accounted for hospital-level clustering and the analysis restricted to continuously enrolling centers was consistent, but secular changes in center composition cannot be fully excluded. Finally, these findings derive from a single national health system. Korea’s pandemic response, post-acute rehabilitation infrastructure, and family-based caregiving norms differ from those of other high-income settings, and the magnitude of the post-2019 inflection may not generalize. Whether a similar inflection occurred elsewhere warrants examination in other registries.

In this 12-year national cohort, a decade of improving outcomes in the oldest-old was followed by deterioration after 2019 despite continued treatment intensification. The change was clearest for functional outcome, for which the age-group-by-year interaction remained significant after adjustment. Because in-hospital treatment and workflow were maintained throughout, sustaining outcome gains in a growing oldest-old population may depend on post-acute care and caregiving, domains that acute stroke registries do not routinely capture.

## Sources of Funding

This research was supported partly by the “Korea National Institute of Health” research project (project No. 2023-ER-1006-02) and a grant of the Korea Health Technology R&D Project through the Korea Health Industry Development Institute (KHIDI), funded by the Ministry of Health & Welfare, Republic of Korea (grant number: HI23C0359).

## Disclosures

Hee-Joon Bae reports grants from Amgen Korea Limited., Celltrion, Dong-A ST, Otsuka Korea, Samjin Pharm, and Takeda Pharmaceuticals Korea Co., Ltd., and personal fees from Amgen Korea, Bayer, Boehringer Ingelheim, Daewoong Pharmaceutical Co., Ltd., Daiichi Sankyo Korea, Eisai Korea, Inc., and Otsuka Korea, outside the submitted work. PBG reports grants from the Havey Institute for Global Health, Northwestern University and NIH Fogarty Institute Center, and support from JLK, American Telephysicians/NeuroX, and Quantal X.

## Data Sharing Statement

Individual deidentified participant data from the CRCS-K-NIH registry that underlie the results reported in this article, together with the data dictionary, study protocol, and statistical analysis plan, will be made available to qualified investigators on reasonable request beginning at the time of publication. Requests, including a methodologically sound research proposal and IRB documentation from the requesting institution, should be directed to the corresponding author (Hee-Joon Bae) and will be reviewed by the CRCS-K-NIH Steering Committee. A signed data use agreement is required, and approved data will be provided in a deidentified format in accordance with the Korean Personal Information Protection Act.

## Supplemental Material

eTables 1-9

eFigures 1-5

## Notes

### Clinical Trial

Not applicable

## References

1. He W, Goodkind D, PaulKowal a, et al. Asia Aging: Demographic, Economic, and Health Transitions. U.S. Government Publishing Office,Washington, DC: U.S. Census Bureau,International Population Reports, 2022.

2. United Nations. Department of E, Social A. World Population Prospects 2024 : Summary of Results. New York: United Nations; 2024.

3. Bejot Y, Duloquin G, Graber M, Garnier L, Mohr S, Giroud M. Current characteristics and early functional outcome of older stroke patients: a population-based study (Dijon Stroke Registry). Age Ageing 2021; 50(3): 898–905.

4. Toyoda K, Yoshimura S, Nakai M, et al. Twenty-Year Change in Severity and Outcome of Ischemic and Hemorrhagic Strokes. JAMA Neurol 2022; 79(1): 61–9.

5. Peng Y, Tan MMC, Ngo L, Alghamry A, Colebourne K, Ranasinghe I. National trends in stroke hospitalisations, mortality, and recurrent stroke in Australia and New Zealand. Sci Rep 2025; 15(1): 42315.

6. Otite FO, Saini V, Sur NB, et al. Ten-Year Trend in Age, Sex, and Racial Disparity in tPA (Alteplase) and Thrombectomy Use Following Stroke in the United States. Stroke 2021; 52(8): 2562–70.

7. Bejot Y. Age Gap between Stroke Patients Included in Randomized Clinical Trials of Acute Revascularization Therapy and Those in Population-Based Studies: A Review. Neuroepidemiology 2023; 57(2): 65–77.

8. van Marum RJ. Underrepresentation of the elderly in clinical trials, time for action. Br J Clin Pharmacol 2020; 86(10): 2014–6.

9. Prabhakaran S, Gonzalez NR, Zachrison KS, et al. 2026 Guideline for the Early Management of Patients With Acute Ischemic Stroke: A Guideline From the American Heart Association/American Stroke Association. Stroke 2026.

10. Kleindorfer DO, Towfighi A, Chaturvedi S, et al. 2021 Guideline for the Prevention of Stroke in Patients With Stroke and Transient Ischemic Attack: A Guideline From the American Heart Association/American Stroke Association. Stroke 2021; 52(7): e364–e467.

11. Goyal M, Menon BK, van Zwam WH, et al. Endovascular thrombectomy after large-vessel ischaemic stroke: a meta-analysis of individual patient data from five randomised trials. Lancet 2016; 387(10029): 1723–31.

12. Ding L, Peng B. Efficacy and safety of dual antiplatelet therapy in the elderly for stroke prevention: a systematic review and meta-analysis. Eur J Neurol 2018; 25(10): 1276–84.

13. Saini V, Guada L, Yavagal DR. Global Epidemiology of Stroke and Access to Acute Ischemic Stroke Interventions. Neurology 2021; 97(20S): S6–S16.

14. D’Cruz M, Banerjee D. ’An invisible human rights crisis’: The marginalization of older adults during the COVID-19 pandemic - An advocacy review. Psychiatry Res 2020; 292: 113369.

15. Nogueira RG, Qureshi MM, Abdalkader M, et al. Global Impact of COVID-19 on Stroke Care and IV Thrombolysis. Neurology 2021; 96(23): e2824–e38.

16. Gabet A, Grave C, Tuppin P, Chatignoux E, Bejot Y, Olie V. Impact of the COVID-19 pandemic and a national lockdown on hospitalizations for stroke and related 30-day mortality in France: A nationwide observational study. Eur J Neurol 2021; 28(10): 3279–88.

17. Katsanos AH, Palaiodimou L, Zand R, et al. Changes in Stroke Hospital Care During the COVID-19 Pandemic. Stroke 2021; 52(11): 3651–60.

18. Bae H-J, Kim JY, Kang J, et al. David G. Sherman Lecture Award: 15-Year Experience of the Nationwide Multicenter Stroke Registry in Korea. Stroke 2022: 101161STROKEAHA122039212.

19. Kim DY, Park TH, Cho Y-J, et al. Contemporary Statistics of Acute Ischemic Stroke and Transient Ischemic Attack in 2021: Insights From the CRCS-K-NIH Registry. J Korean Méd Sci 2023; 39(34): e278.

20. Jr HPA, Bendixen BH, Kappelle LJ, et al. Classification of subtype of acute ischemic stroke. Definitions for use in a multicenter clinical trial. TOAST. Trial of Org 10172 in Acute Stroke Treatment. Stroke 2018; 24(1): 35–41.

21. Chaisinanunkul N, Adeoye O, Lewis RJ, et al. Adopting a Patient-Centered Approach to Primary Outcome Analysis of Acute Stroke Trials Using a Utility-Weighted Modified Rankin Scale. Stroke 2015; 46(8): 2238–43.

22. Adcock AK, Schwamm LH, Smith EE, et al. Trends in Use, Outcomes, and Disparities in Endovascular Thrombectomy in US Patients With Stroke Aged 80 Years and Older Compared With Younger Patients. JAMA Netw Open 2022; 5(6): e2215869.

23. Andersen KK, Andersen ZJ, Olsen TS. Predictors of early and late case-fatality in a nationwide Danish study of 26,818 patients with first-ever ischemic stroke. Stroke 2011; 42(10): 2806–12.

24. Elm Ev, Altman DG, Egger M, et al. The Strengthening the Reporting of Observational Studies in Epidemiology (STROBE) statement: guidelines for reporting observational studies. The Lancet 2007; 370(9596): 1453–7.

25. Xian Y, Li S, Jiang T, et al. Twenty Years of Sustained Improvement in Quality of Care and Outcomes for Patients Hospitalized With Stroke or Transient Ischemic Attack: Data From The Get With The Guidelines-Stroke Program. Stroke 2024.

26. Conrad Drescher FB, Teresa Ullberg, Mats Pihlsgård, Bo Norrving, Jesper Petersson. Temporal changes in functional outcome and case-fatality after ischaemic stroke and intracerebral haemorrhage in Sweden 2010–2019:an observational study fro m the Swedish Stroke Register (Riksstroke). European Stroke Journal 2026; 11(1): 1–11.

27. Urfy M, Mir MT. Trends and Disparities in In-Hospital Mortality Among Ischemic Stroke Patients During the COVID-19 Era: A Nationwide Study (2016–2022). J Stroke Cerebrovasc Dis 2025; 34(8): 108367.

28. Xian Y, Xu H, Matsouaka R, et al. Analysis of Prescriptions for Dual Antiplatelet Therapy After Acute Ischemic Stroke. JAMA Netw Open 2022; 5(7): e2224157.

29. Johnston SC, Easton JD, Farrant M, et al. Clopidogrel and Aspirin in Acute Ischemic Stroke and High-Risk TIA. New Engl J Med 2018; 379(3): 215–25.

30. Wang Y, Wang Y, Zhao X, et al. Clopidogrel with Aspirin in Acute Minor Stroke or Transient Ischemic Attack. New Engl J Med 2013; 369(1): 11–9.

31. Chen H, Jindal G, Miller TR, Gandhi D, Chaturvedi S. Stroke Thrombectomy in the Elderly: Efficacy, Safety, and Special Considerations. Stroke Vasc Interv Neurol 2023; 3(4): e000634.

32. Ko D, Lin KJ, Bessette LG, et al. Trends in Use of Oral Anticoagulants in Older Adults With Newly Diagnosed Atrial Fibrillation, 2010-2020. JAMA Netw Open 2022; 5(11): e2242964.

33. Jian Y, Zhao L, Jia B, et al. Direct versus Bridging Mechanical Thrombectomy in Elderly Patients with Acute Large Vessel Occlusion: A Multicenter Cohort Study. Clin Interv Aging 2021; 16: 1265–74.

34. Lin DSH, Lo HY, Huang KC, Lin TT, Lee JK. Efficacy and Safety of Direct Oral Anticoagulants for Stroke Prevention in Older Patients With Atrial Fibrillation: A Network Meta-Analysis of Randomized Controlled Trials. J Am Hear Assoc: Cardiovasc Cerebrovasc Dis 2023; 12(23): e030380.

35. Silverio A, Cancro FP, Esposito L, et al. Secondary Cardiovascular Prevention after Acute Coronary Syndrome: Emerging Risk Factors and Novel Therapeutic Targets. J Clin Med 2023; 12(6).

36. Ganesh A, Luengo-Fernandez R, Wharton RM, et al. Time Course of Evolution of Disability and Cause-Specific Mortality After Ischemic Stroke: Implications for Trial Design. J Am Heart Assoc 2017; 6(6).

37. Thau L, Siegal T, Heslin ME, et al. Decline in Rehab Transfers Among Rehab-Eligible Stroke Patients During the COVID-19 Pandemic. J Stroke Cerebrovasc Dis 2021; 30(8): 105857.

38. Mayer-Suess L, Ter Telgte A, Praxmarer S, et al. Stroke Care Pathway ensures high-quality stroke management in the COVID-19 pandemic. Sci Rep 2023; 13(1): 5587.

39. Siegler JE, Abdalkader M, Michel P, Nguyen TN. Therapeutic Trends of Cerebrovascular Disease during the COVID-19 Pandemic and Future Perspectives. J Stroke 2022; 24(2): 179–88.

40. Ryu SI, Park YH, Kim J, et al. Impact of COVID-19 on the social relationships and mental health of older adults living alone: A two-year prospective cohort study. PLoS One 2022; 17(7): e0270260.

41. Lee G, Kim C. Social isolation and mental well-being among Korean older adults: a focus on living arrangements. Front Public Health 2024; 12: 1390459.

42. Hu B, Shin P, Han EJ, Rhee Y. Projecting Informal Care Demand among Older Koreans between 2020 and 2067. Int J Environ Res Public Health 2022; 19(11).

43. KS P. Consecrating or Desecrating Filial Piety?: Korean Elder Care and the Politics of Family Support. Journal of Asian Sociology 2013; 42(2): 287–308.

44. Kim E, Kim JY, Moon KM, et al. One-year mortality and associated factors in older hospitalized COVID-19 survivors: a Nationwide Cohort Study in Korea. Sci Rep 2024; 14(1): 24889.

45. Busis NA, Shanafelt TD, Keran CM, et al. Author response: Burnout, career satisfaction, and well-being among US neurologists in 2016. Neurology 2017; 89(15): 1650–1.

46. Dall TM, Storm MV, Chakrabarti R, et al. Supply and demand analysis of the current and future US neurology workforce. Neurology 2013; 81(5): 470–8.

47. Yoon JH, Kwon IH, Park HW. The South Korean health-care system in crisis. The Lancet 2024; 403(10444): 2589.

48. Downer MB, Luengo-Fernandez R, Binney LE, et al. Association of multimorbidity with mortality after stroke stratified by age, severity, etiology, and prior disability. Int J Stroke 2024; 19(3): 348–58.

49. De Cola MC, Ielo A, Corallo F, et al. Development of a Set of Indicators for Measuring and Improving Quality of Rehabilitation Care after Ischemic Stroke. Healthcare (Basel) 2023; 11(14).

50. Park TH, Hong KS, Cho YJ, et al. Temporal Trends in Stroke Management and Outcomes Between 2011 and 2020 in South Korea: Results From a Nationwide Multicenter Registry. J Am Hear Assoc: Cardiovasc Cerebrovasc Dis 2025; 14(5): e035218.

51. Sattar N, Preiss D, Murray HM, et al. Statins and risk of incident diabetes: a collaborative meta-analysis of randomised statin trials. The Lancet 2010; 375(9716): 735–42.

